# The first thousands of cases of coronavirus disease 2019 (COVID-19) in Algeria: some risk factors

**DOI:** 10.1101/2020.08.17.20176396

**Authors:** Salima Taleb, Marwa Boussakta

## Abstract

**Objectives:** Providing valuable information on the prevalence of Covid-19 is a crucial step to improve and accentuate the disease surveillance and prevention system as this can limit the spread of the virus.

**Methods:** COVID-19 is caused by the SARS-CoV-2 virus. It is essential to understand the epidemiological characteristics of the first cases in each country. The purpose of this study is to describe the geographic distribution and some risk factors in the first thousands of cases in Algeria. This descriptive study was carried out to examine recent data published by public health institutions in Algeria, websites and the world health organization.

**Results:** The 8306 cases of COVID-19 have been confirmed in Algeria. By sex, men with 55.76% predominate, the most affected age group was 25 to 49 years old (41.1%), 600 cases of death were reported, subjects aged over 60 years are the most likely to die from COVID-19. Most of the confirmed subjects came from the cities of Blida and Algiers. All cases are human-to-human transmissions.

**Conclusion:** The COVID-19 pandemic has highlighted the lack of dical equipment in Algeria and in all countries of the world. This requires better management of the health sector on an international scale.

## Introduction

Since December 2019, a number of cases of pneumonia of unknown origin have appeared in Wuhan, China’s Hubei Province [1]. This disease has spread rapidly around the world in many countries across 6 continents. According to the Minister of Health, the national incidence of Covid-19 in Algeria is currently 84 per 100000. On January 3, 2020, a new member of the coronavirus family (with enveloped RNA) was detected in broncho-alveolar lavage fluid from a patient in Wuhan and subsequently identified and confirmed as the cause of this disease by the Chinese Center for Disease Control and Prevention (CDC) [2–4]. On January 7, 2020, the World Health Organization (WHO) named this new 2019 coronavirus, (SARS-CoV-2). SARS-CoV-2 shares 80% homology with SARS-CoV and 96% similarities to a bat virus. SARS-CoV-2 has an ACE2 receptor, similar to SARS-CoV and NL 63 coronavirus, capable of multiplying in the respiratory epithelium [5] (MSPRH, DGPPS).

In February 2020, WHO names this new disease associated with coronavirus 2019, (COVID-19). The spread of SARS-CoV-2 has drawn worldwide attention and the WHO declared COVID-19 a global pandemic [6] as of March 11, 2020 [7]. The most common symptoms of COVID-19 are fever, dry cough, and fatigue. Some patients experience pain, nasal congestion, sore throat or diarrhea. These symptoms generally resemble those of a simple flu. Some carriers of this virus have only very discreet symptoms [8]. As of May 24, 2020, 5380774 cases have been confirmed worldwide with no less than 343880 reported deaths (MSPRH [9].

According to several theories, the reservoir of the virus is probably animal. Even if SARSCoV-2 is very close to a virus detected in a bat, the animal responsible for transmission to humans has not yet been identified with certainty [7]. As a new pandemic, cases have been reported in several other countries in Asia, Europe, America, Australia and Africa [10].

Transmission can occur from animals to humans as well as human-to-human transmission by aerosol and droplets. There is also a possibility of oral fecal transmission, since nucleic acids have been found in fecal and anal samples from confirmed patients [11].

Currently, the best diagnostic tool available is the reverse transcription polymerase chain reaction (RT-PCR), unfortunately, it has a high false positive rate, so thin section chest tomography has an important role in diagnosis of this disease [12, 13].

Despite the world spread, the epidemiological and clinical patterns of COVID-19 are not yet clear, particularly in children, pregnant women and the obese. Due to these gaps in the information published by several national and world organizations, we will present in this study the first prevalence’s of COVID-19 according to some risk factors in Algeria.

## Methods

### Study population

Since February 25, 2020, until May 24, all confirmed patients with coronavirus disease 2019 (COVID-19) in Algeria were included in this study. The diagnosis of COVID-19 was based on guidelines from the World Health Organization.

### Course of the study

This descriptive study was conducted to review key information from the epidemiological bulletins of the Ministry of Population Health and Hospital Reform (MSPRH) and reports from the World Health Organization (WHO).

MSPRH and WHO data were used to present descriptive results for categorical variables. In this study, we collected data published from February 25, 2020 to May 24, 2020. We conducted a retrospective study on the epidemiological characteristics of 8306 COVID-19 patients of all ages (3675 female sexes and 4631 male sexes).

### COVID-19 Confirmation

The presence of SARS-CoV-2 was detected by the real-time reverse transcription polymerase chain reaction (RT-PCR) and simple PCR method, carried out by the Pasteur Institute of Algeria (IPA) and its annexes in Constantine, Oran and in other regions of the country (El Oued, TiziOuzou …). Once the biological and virological results have been obtained, the diagnosis is completed by an X-ray examination (chest scanner) for confirmation in order to decide on the course of action to follow.

### Statistical

Data were entered and processed by Excel 2007 software. Results are presented as number and frequency.

## Results

Since February 25, 2020, 8306 cases of COVID-19 have been confirmed in Algeria. The number of patients increased from 1 confirmed case on February 25, 2020 to 8306 confirmed cases on May 24, 2020 (Figure 1). Depending on the place of residence, most of the infected subjects came from the city of Blida (13%), Algiers (11%), Oran (6%), Setif (6%) and Constantine (5%). The prevalence of COVID-19 by city is presented in figure 2. The age group most affected by COVID-19 is 25–49 years old with 41.1% followed by people over 60 years old with 30,3%, the distribution of the different age groups according to the prevalence of the disease is illustrated in figure 3. Taking sex into account, men are more affected than women (55.76% vs 44.24%), figure 4. Among the patients with COVID-19 who benefited from the chloroquine-based treatment protocol, 4578 were cured. From the onset of the disease until May 24, 2020, 600 patients have died (Figure 5). The prevalence of deaths by age group is illustrated in figures 7. The number of COVID-19 cases by country until May 24, 2020 is presented in Figure 6, in this figure, the number highest case was observed in the United States of America with 1592,599 cases.

**Figure 1:**
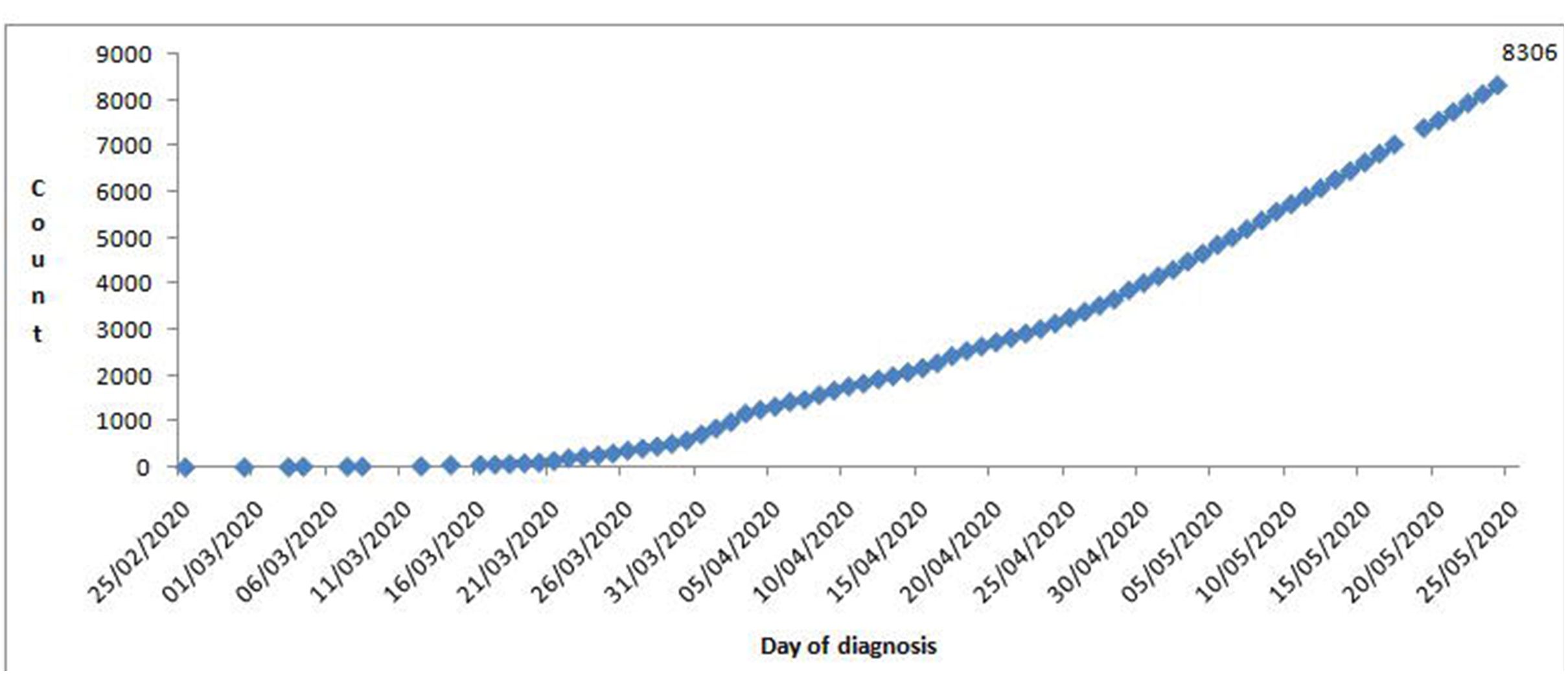
Evolution of the number of cases of COVID-19 in Algeria according to the day of diagnosis (Source: from the Ministry of Health until May 24, 2020)

**Figure 2:**
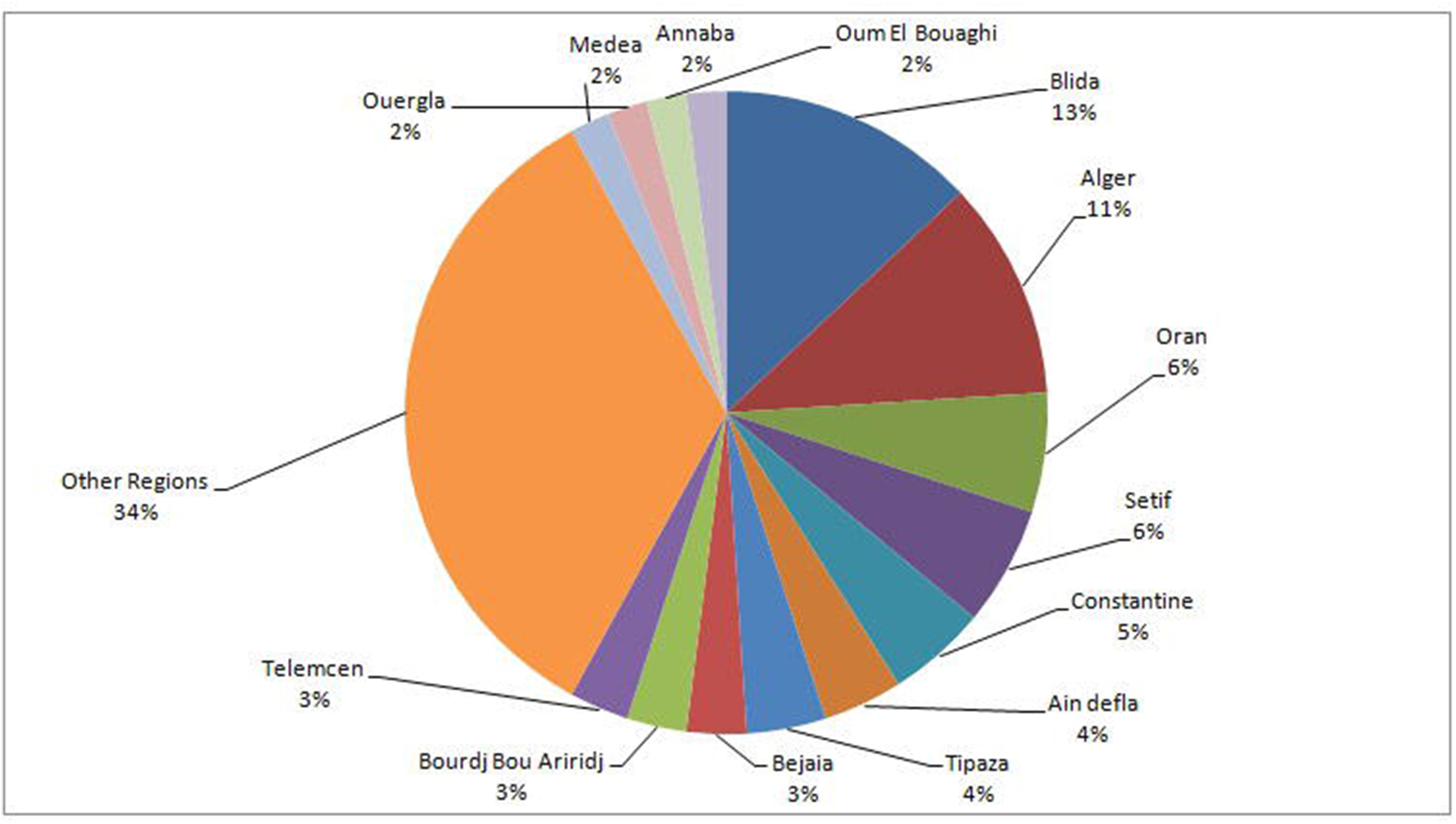
Prevalence of COVID-19 cases confirmed by city until May 24, 2020 in Algeria.

(Source: Ministry of Health)

**Figure 3:**
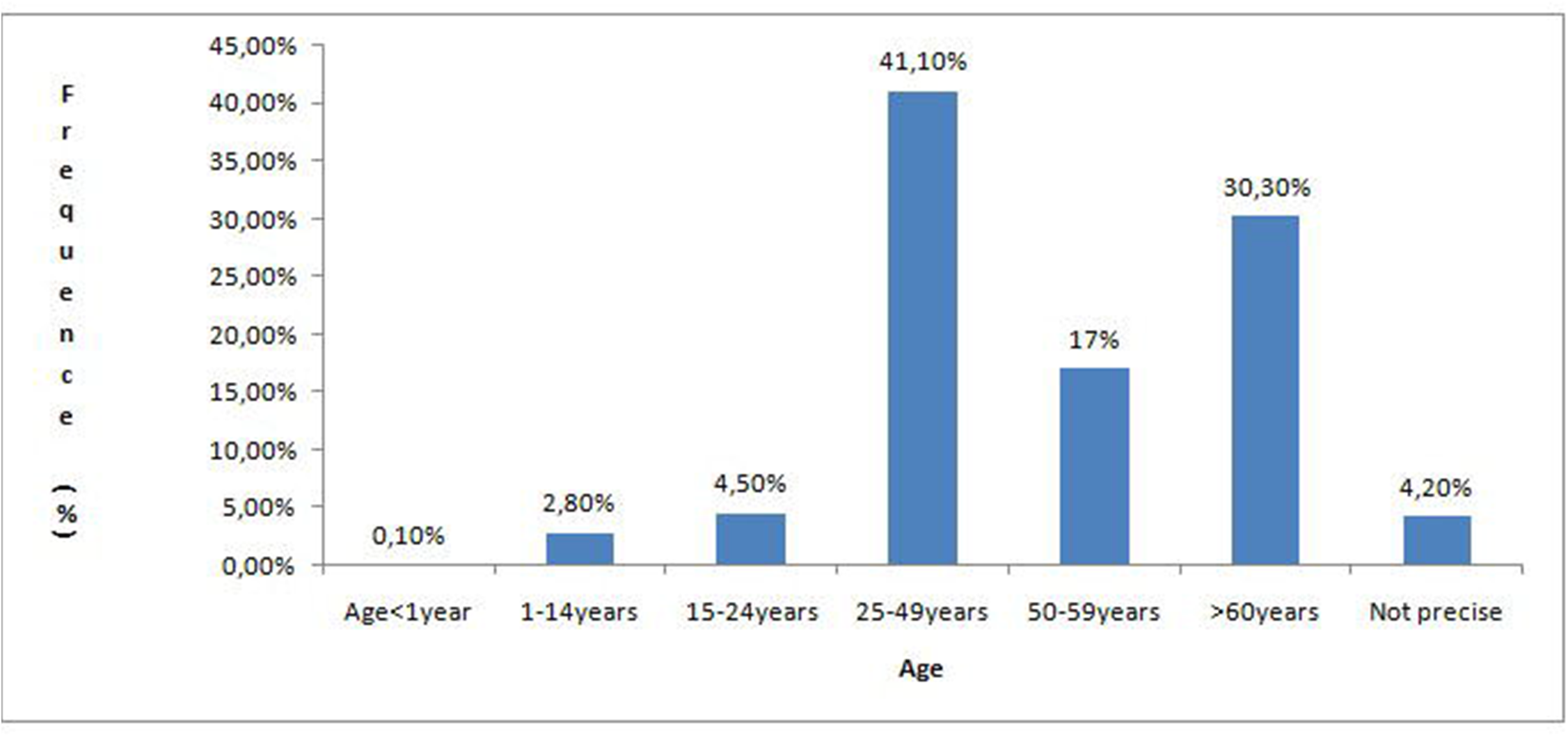
Prevalence of COVID-19 confirmed cases by age group until May 24, 2020 in Algeria.

(Source from the Ministry of Health until May 24, 2020)

**Figure 4:**
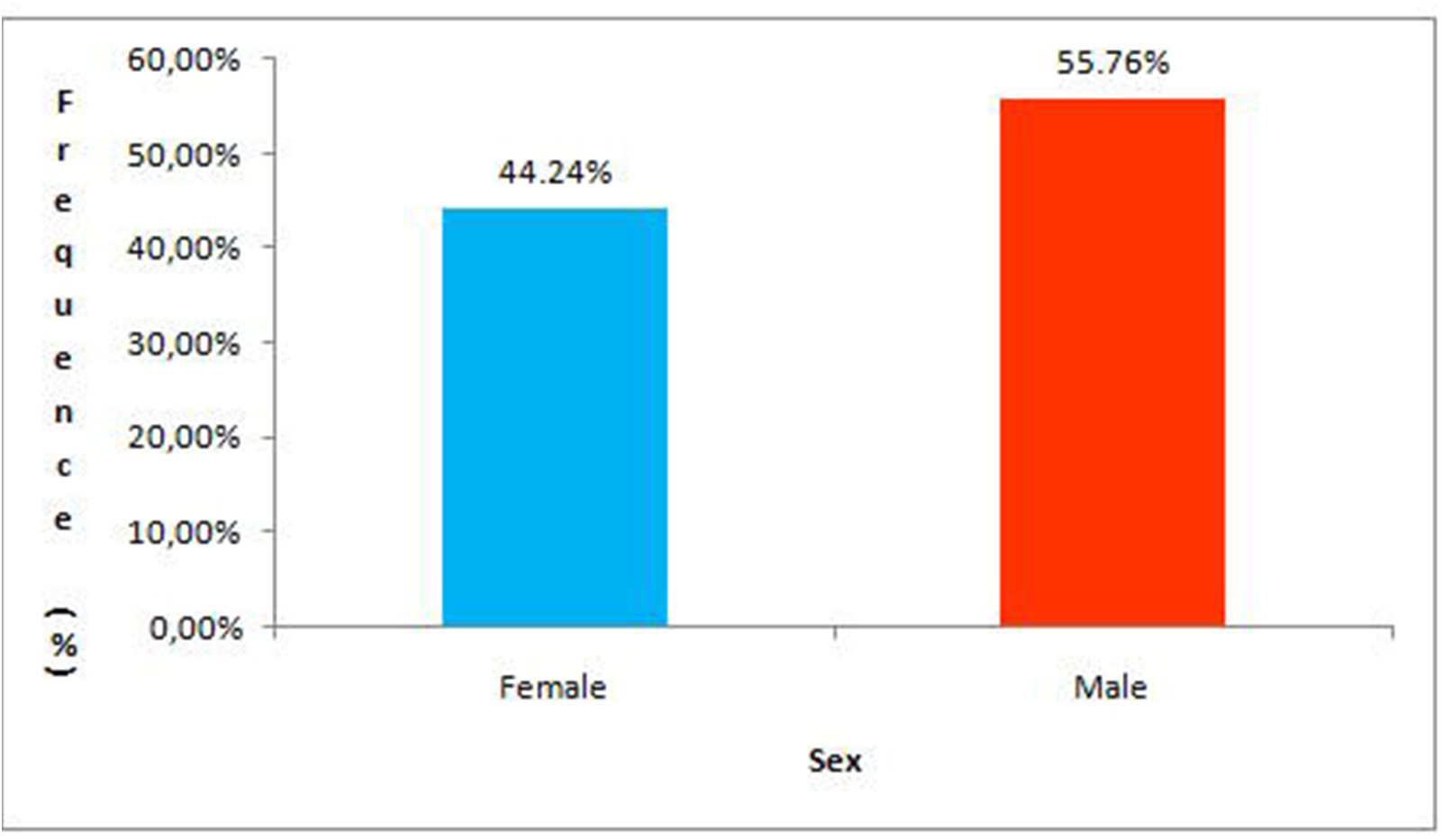
Prevalence of COVID-19 confirmed cases by sex until May 24, 2020 in Algeria.

(Source: Ministry of Health).

**Figure 5:**
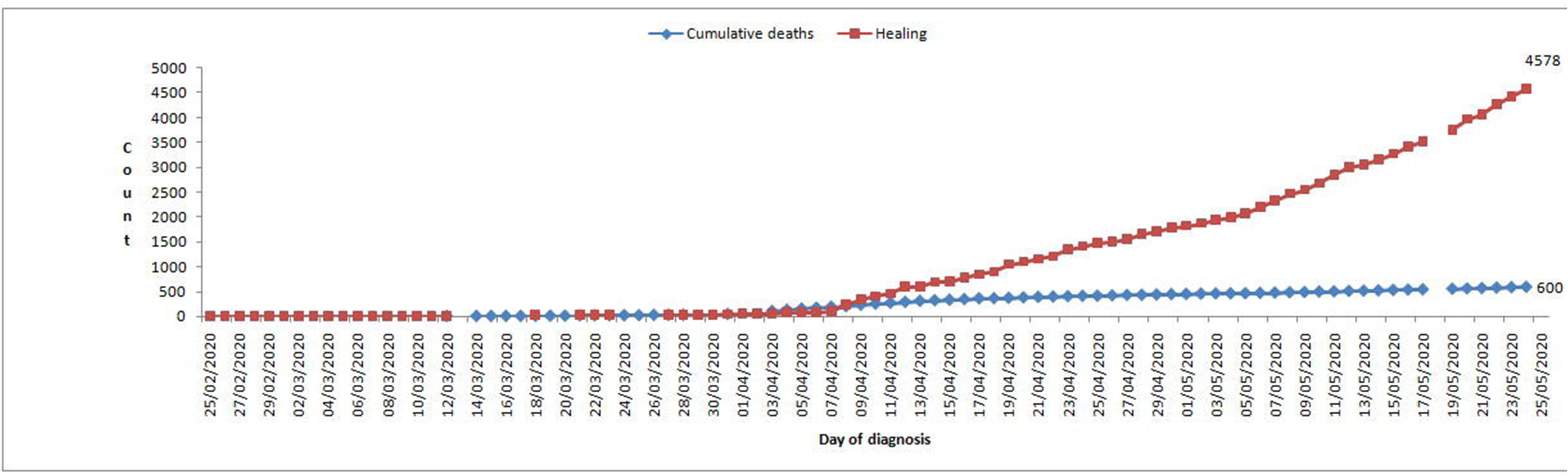
Prevalence of cases of death and recovery until May 24, 2020 in Algeria.

(Source: Ministry of Health).

**Figure 6:**
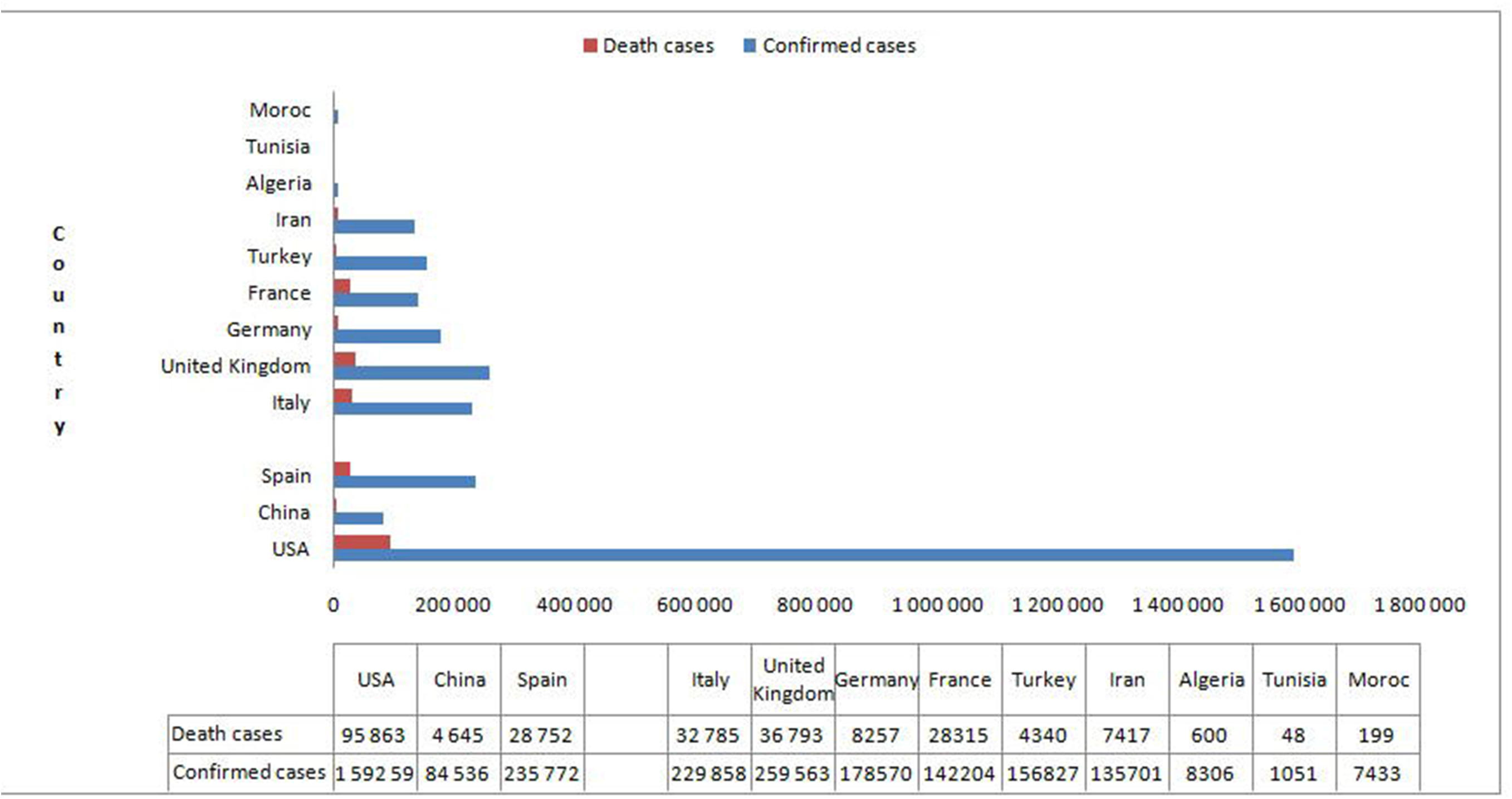
Number of confirmed cases and deaths of COVID-19 by country since the onset of the disease until May 24, 2020 (Source WHO: http://www.who.int ›situation reports.

(Source: Ministry of Health).

**Figure 7:**
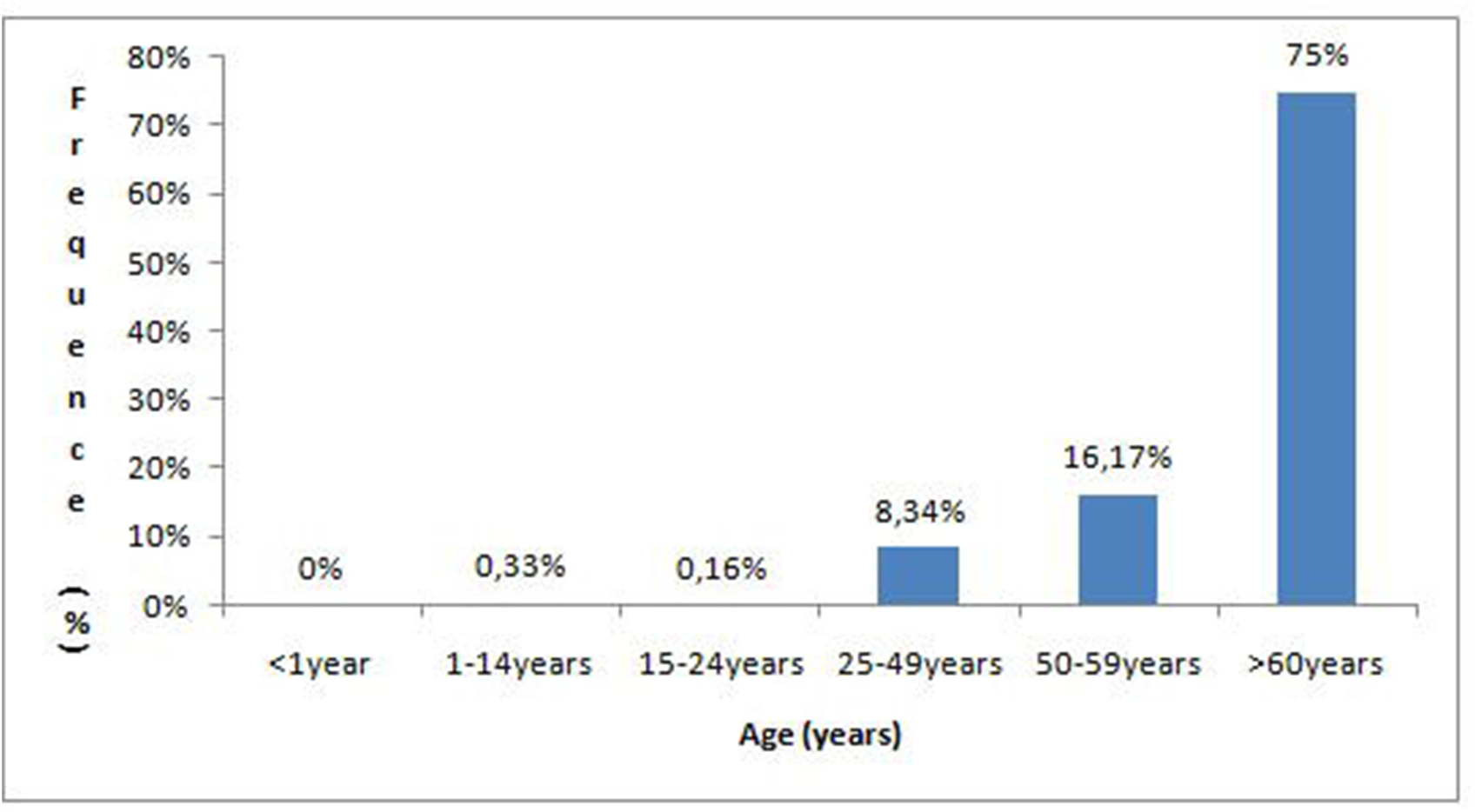
Prevalence of total deaths from COVID-19 by age group until May 24, 2020 in Algeria.

(Source: Ministry of Health)

## Discussion

This is the first retrospective study on the prevalence of COVID-19 in Algeria according to certain risk factors. This study brought the number of confirmed cases of COVID-19 since its appearance in Algeria on February 25, 2020 until May 24, 2020. Since all these confirmed cases whether adults or children were likely to be exposed to family members and / or other people in work, supermarkets, public transport … reached of COVID-19 that leads us to say that the transmission is clearly person-to-person. Evidence that supported this route of transmission has been reported by other studies [14, 15, 16, and 17]. This mode of transmission has made this disease a very serious pandemic, although it is not as severe as that of other diseases caused by other viruses. COVID-19, not only impacted health systems around the world, but also disrupted daily life and economy in several parts of the world, it also caused great panic, which was intensified by the media and the lack of centralized dissemination of reliable information [18].

As in many countries around the world, the disease appeared in the most populous areas such as Blida and Algiers, and then it spread to all the cities of the country. This has been reported in other studies [18]. Most of the cases affected in this study are male. The statistics of the corona virus are still too incomplete to decide, but the same result has been observed in Colombia, China and Italy through recently published studies [18, 19, 20]. We believe that this predominance of men is due to the differences between sex in the way of life and the prevalence of certain diseases. In Algeria, we are now witnessing a real epidemiological transition marked by changes in lifestyle, with smoking in epidemic upsurge and chronic and non-communicable diseases which have taken a growing place and currently constitute major public health problems in Algeria. In Algeria in 2016, 30.4% of men smoke against only 0.7% of women [21]. Mortality from cardiovascular disease, cancer and diabetes was estimated at 14.2% in 2016 [21]. Information has shown that 22% of Algerian women are exposed to heart attacks in Algeria [22]. This is what may have implications for mortality from the COVID-19 epidemic. The same observations have been reported by Chinese researchers [23, 24]. The data we have collected does not contain any information on the health status of the patients, which leaves these observations unlikely and requires further clarification. The age group most affected by COVID-19 in Algeria is 25–49 years old. This is the most active age group in the population and often comes into contact with other people. This is what leads us to assume that the transmission is person-to-person. This mode of transmission is confirmed in the WHO report [10]. Children and adolescents are the least affected because they are often controlled by parents. In a recently published Chinese study, the authors tell us that children affected by COVID-19 do not show severe clinical signs as in adults. Unfortunately in our study we cannot give more details since we do not have the clinical characteristics of the population studied. Of 8,306 confirmed cases, 600 (7.22%) died of which 450 (75%) are over 60 years old. As in other studies [25], the first factor associated with C0VID-19 mortality is unsurprisingly age. According to the same study, those over 80 are 12 times more likely to die than the 50–60 age group. The risk is even greater when the results have been compared with those 40–50 years old. For our study, we cannot give more details due to lack of information. Despite the spread of COVID-19 which continues to increase day by day as shown by the results of this study, the number of recoveries remains a positive point despite the lack of an effective treatment for this disease.

The number of cases in Algeria has continued to increase since May 24, 2020, which makes the situation a bit critical. Failure to comply with confinement rules does not allow the Algerian authorities to control the situation as they plan.

## Conclusion

We report in this study early data on the prevalence of COVID-19 in Algeria and in the world using the data available on the website of the Ministry of Population Health and Hospital Reform of Algeria as well as the data of the first reports published by the World Health Organization on COVID-19. It is important to note that the statistical estimates available to date are preliminary. They must be supplemented by large-scale epidemiological studies, which so far have been lacking, whether in Algeria, China, Italy, France or elsewhere in the world. Additional work is planned on this problematic. However, it should be noted that the results of this study showed that the transmission is human-to-human and that the oldest people and men are the most affected. Finally, we mention that the public health measures implemented by different countries show that certain changes must be implemented.

## Data Availability

All data used in this study are available

https://www.sante.gov.dz

https://www.who.int

## Declaration of interests

The authors declare that they have no conflicts of interest in relation to this article.

### Ethics Statement

All procedures performed in studies involving human participants were in accordance with the ethical standards of the institutional research committee (NCC2014–0068) and with the 1964 Helsinki declaration and its later amendments or comparable ethical standards.

## Supplementary Materials

“None.”

## Author Contributions

Concept and design, Data curation, Formal analysis and interpretation of data, Drafting of the manuscript, Methodology, Writing – review & editing: Salima TALEB et Marwa BOUSSAKTA.

## Acknowledgment

The authors thank the MSPRH for its assistance. They thank the World Health Organization for its help, for making all of the data that went into this article available to us.

